# A community-based structured birth planning intervention to increase skilled delivery and decrease neonatal mortality: a cluster randomised controlled trial in Pakistan

**DOI:** 10.64898/2026.09.29.26364292

**Authors:** Muhammad Amir Khan, John D. Walley, Wajiha Javed, Nida Khan, Maqsood Ahmad, Shaheer Ellahi Khan, Joseph Paul Hicks, Rebecca King, Robert West

## Abstract

**Background:** We developed and evaluated a behaviour change intervention to improve health outcomes for pregnant women and neonates in rural Pakistan.

**Methods:** We used a parallel, three-arm, unblinded, cluster randomised controlled trial design. Eligible clusters were upgraded health facilities plus all associated community health workers (CHWs) and catchment populations. Randomisation was by computer program (1:1:1 ratio). All pregnant women were eligible. Control CHWs provided usual care to pregnant women. Arm 1 CHWs provided structured birth planning, including birth preparedness and complication readiness planning, and a voucher allowing free travel to access emergency obstetric/neonatal care. Arm 2 CHWs provided structured planning alone. Co-primary outcomes were skilled (doctors and midwifery-trained cadres) birth attendance and neonatal mortality (NNM) at 28 days.

**Findings:** Between November 2011 to May 2012, respectively in arms 1, 2 and the control 21,317, 19,267 and 18,867 women were recruited, and 21,214, 19,125 and 18,812 were followed-up and analysed. Vouchers were almost never used to reclaim travel costs. Skilled attendance was 64% (12,045/18,812) in the control arm, 72.3% (15,335/21,214) (adjusted odds ratio [aOR] 1.56 [95% CI: 0.93, 2.62; p = 0.21]) in arm 1 and 78.1% (14,944/19,125) (aOR 2.16 [95% CI: 1.29, 3.62; p = 0.007]) in arm 2. NNM was 13.4 (per 1,000 live births) in the control arm, 45 (aOR 2.32 [95% CI: 1.48, 3.63; p < 0.001]) in arm 1 and 16.2 (aOR 1.36 [95% CI: 0.84, 2.21; p = 0.21]) in arm 2.

**Conclusions:** Birth planning had inconsistent effects on skilled attendance and NNM, including apparently negative effects on NNM, with concerns about the quality of midwifery care. However, the control arm and arm 2 NNM was improbably low for this setting, and bias in the outcome may explain the observed differences. The travel voucher was little used.

**Trial Registration:** ISRCTN 86264432 (DOI 10.1186/ISRCTN86264432).

## Introduction

Historically Pakistan has had one of the world’s highest rates of neonatal and maternal mortality, and in 2015 these rates remained very high at 45.5 per 1000 live births and 178 per 100,000 live births respectively. A major cause of these poor neonatal and maternal reproductive outcomes is the low rate of deliveries by skilled birth attendants (just 52% in 2013) [1]. Since 2006 Pakistan’s National Programme for Family Planning and Primary Health Care (commonly referred to as the Maternal Neonatal and Child Health (MNCH) Programme) has aimed to address these issues primarily by promoting skilled birth attendance through birth preparedness, strengthening the primary and secondary levels of healthcare by ensuring quality comprehensive emergency obstetric and neonatal care (EmONC) (able to provide medications, manually remove placenta and retained products, perform assisted vaginal delivery and neonatal resuscitation) is provided at rural health centres, and comprehensive EmONC (basic plus caesarean section and blood transfusion) is provided at hospitals, and by promoting prenatal, postnatal and new born care.

In Pakistan a large and well-functioning network of around 110,000 community health care workers (CHWs), known as lady health workers, provide care for around 250 households each. As well as providing antenatal and postnatal care these CHWs act as ‘gatekeepers’, promoting skilled birth attendance at delivery and use of EmONC (where necessary) via birth-preparedness planning (informed selection and organisation of a skilled birth attendant for delivery) and complication-readiness planning (informed selection of a facility providing EmONC plus pre-arranged travel plans if needed) with pregnant women and their family members. However, there are concerns over how effective CHWs are in this role as gatekeepers, and they currently lack structured guidelines and tools to effectively counsel pregnant women and their families, to facilitate their birth-preparedness and complication-readiness planning. In addition, although more generally it is clear that, amongst other key factors, increasing the proportion of deliveries by skilled birth attendants and improving access to quality antenatal and postnatal care can substantially improve neonatal and maternal reproductive outcomes, there is little robust evidence on the effectiveness of birth-preparedness planning [2].

We therefore developed two interventions that were delivered by CHWs: 1) structured birth planning, which involved CHWs helping pregnant women and their families develop birth-preparedness and birth-complication-readiness plans using specifically designed structured guidelines and information tools, plus a travel voucher that would allow pregnant women to claim back transportation costs if they needed to access EmONC services during birth, and 2) structured birth planning without a travel voucher. We hypothesised that our structured birth planning intervention would enhance the ability of CHWs to increase pregnant women’s use of skilled birth attendants, and that providing pregnant women with the travel voucher would increase their willingness to rapidly seek EmONC care if required, so that both interventions would reduce neonatal and maternal mortality. Here we present the results of a cluster randomised trial evaluating the effectiveness of these interventions.

## Methods

The CONSORT for cRCTs checklist is included as a supplementary material (S1 CONSORT Checklist), and the protocol for this trial has been previously published [3].

### Study design, setting and participants

The study was done across three districts (Chiniot, Jhang and Khanewal) in Punjab province, Pakistan. We and used a parallel three-arm, cluster-randomised controlled superiority design to compare each intervention arm to a control arm. We defined a cluster as either a sub-district (tehsil) hospital or a rural health centre (RHC) plus all the CHWs linked to that health facility or its associated basic health units (BHUs), along with all the pregnant women covered by those CHWs (Table 1). Clusters were eligible if 1) the health facility was capable of providing comprehensive EmONC and had at least one qualified doctor present, and 2) the health facility had an associated functioning network of CHWs. Across the three districts there were 26 potential cluster health facilities in total: 6 in Chiniot, 12 in Jhang and 8 in Khanewal.

**Table 1.** Health workers and their facilities/place of work involved in trial.

| **Health worker and general role** | **Facility/place of work** | **Delivery care role** |
| --- | --- | --- |
| Traditional birth attendant (Dai): an unskilled midwife. | Clients' home. | Traditional midwife, normal deliveries only. |
| Lady health worker: a community health worker (CHW). | Work in the community, meeting clients at the client's home or their own home. Associated with an RHC, BHU or sub-district hospital for supervision by lady health supervisors and lady health visitors based at one of these facilities. | Do not assist with delivery, but involved with antenatal and postnatal care of mothers and children, and delivering the trial interventions to women. |
| Lady health visitor: a midwife/maternal and child care provider. Also provide technical supervision and resupply services for CHWs. | Work at a RHC, BHU or sub-district hospital, but may in addition also work from their own private clinic. | Midwife, including normal and instrumental (via forceps) delivery, and also provide antenatal and postnatal care for mothers and children. |
| Lady health supervisor: a monitoring and supervisory cadre for CHWs (usually supervise around 20 CHWs each). | Employed and based in the district by the LHW programme. | No direct delivery care role. |
| Community midwife (CMW): a new cadre of community-based midwives created under the MNCH Programme. | Own home or clients' home. | Midwife, normal deliveries only. |
| Doctor: a fully trained medical doctor. | Sub-district hospital, RHC or BHU. | Normal, instrumental deliveries (forceps/vacuum-assisted) and EmONC caesarean deliveries. |

We chose a cluster randomised design because it would not have been feasible to have CHWs alter their care practices depending on a patient’s treatment allocation, and also to minimise the possibility of contamination between women receiving different interventions. We sought informed communal consent from clusters prior to randomisation. At each of the clusters there was a formal meeting with community representatives. The health centre staff explained the activities in the intervention and trial, and then sought their written informed consent. The study used anonymised data from the available routine records maintained by the CHW.

### Randomisation, recruitment and masking

We decided all 26 clusters were eligible. Based on our required sample size we randomly selected 21 clusters, and obtained communal consent from them all. The trial statistician then randomly allocated all 21 clusters in a 1:1:1 ratio between the three treatment arms using a computer program. During the recruitment phase all consenting pregnant women receiving care from CHWs within the cluster catchment populations were recruited by their CHWs. It was not possible to mask the treatment allocation from the CHWs given their involvement in the interventions, and it was not possible to mask treatment allocation from pregnant women.

### Procedures

We explicitly designed the intervention to be replicable under routine health service provision, with modest external inputs, and along with provincial MNCH experts we drafted the limited changes required to the existing service and system. We compared two interventions, delivered by CHWs, to the existing CHW-led care of pregnant women: structured birth planning plus a voucher allowing free travel to an EmONC facility (SBP and travel voucher), and structured birth planning alone (SBP alone). Within communities CHWs serve about 1,000 women, and as part of their routine duties they are responsible for identifying and registering all new pregnancies, as well as providing basic antenatal care to registered pregnancies, encouraging women to attend skilled antenatal examinations at their nearest health facility, promoting skilled birth attendance, preparing women and families for possible birth complications, and reporting registered pregnancies and their eventual outcomes on a monthly basis through. In their work they are supervised and monitored by a lady health supervisor, employed by the MNCH Programme, as well as by a lady health visitor, who resupplies them with medications and equipment and provides technical support (Table 1).

Prior to the implementation of the intervention all lady health visitors, lady health supervisors and CHWs in the 21 recruited clusters were enlisted. Then, with the support of the district offices, the enlisted lady healthy visitors and lady health supervisors were trained by research staff on a specially designed set of care reporting and monitoring activities to support the implementation and running of the intervention and data collection. Then, with the support of the respective sub-district hospital/RHC staff, the enlisted lady health visitors in the 14 intervention clusters were trained by research staff on a specially designed programme to deliver the intervention components, including conducting structured birth planning, and mapping skilled birth attendants, transport service providers and potential suitable EmONC facilities within their cluster catchment areas. Once trained the lady health visitors then trained the CHWs within the intervention clusters on these activities during the CHWs routine monthly meetings.

CHWs in the intervention arms then mapped all the skilled birth attendants (including public and private midwives and doctors), suitable EmONC facilities (providing basic or comprehensive EmONC) and potential travel providers (to EmONC facilities) in their catchment populations. With research staff support the lady health visitors and lady health supervisors then checked these data for completeness and correctness, and created directories of the mapped services that were then used by CHWs in the intervention arms during structured birth planning sessions.

The structured birth planning intervention was motivated by the relatively low use of available skilled birth attendants and suitable health facilities, as well as concerns around how well and frequently women and family members recognise danger of signs of birth complications and act appropriately (i.e. access EmONC). We therefore aimed to increase informed access to and use of safe birthing services (and EmONC where necessary) for women via this intervention. Structured birth planning therefore included structured birth-preparedness and complication-readiness counselling, which were provided to consenting women in both intervention arms by CHWs during their routine visits, following the registration of a pregnancy. During a structured birth planning session in a clients’ home, CHWs used the directory to help clients select a skilled birth attendant from those available locally, as well as a place of delivery, which was chosen based on careful consideration of the options available (including the ability to access EmONC if needed), with input from family members planning to be present during the delivery, and following communication with their selected skilled birth provider.

They also provided pregnant women and their family members with information on a range of pregnancy and birth related issues focused on birth-preparedness and complication-readiness. A specially designed illustrated education tool (flipchart) was developed to help CHWs simply and effectively deliver this counselling information. Counselling on birth-preparedness covered the following areas: 1) paying for services (e.g. through savings, a personal loan or selling a disposable asset), 2) making a safe delivery kit (a list of consumables required, and arranged in-advance, for hygienic delivery), and 3) pre-delivery communication with the selected birth attendant to discuss and agree on important service details including how they can communicate, the cost and payment method. Counselling on complication-readiness covered the following areas: 1) recognising the danger signs of birth complications, indicating the need for EmONC services, 2)pre-identifying a MNCH Programme endorsed facility offering comprehensive EmONC services, and 3) arranging, in-advance, transport to the chosen EmONC facility, including selecting a transport service provider with a known type of transport, and contacting them in advance, directly or via mobile phone, to discuss their availability, how they can communicate, the cost and payment method. In addition to this intervention-specific information, the CHWs also provided pregnant women with a range of routinely provided information on antenatal care (relating to diet, rest, hygiene and use of iron tablets) and postnatal care (relating to breastfeeding, diet and family planning).

At the end of a counselling session CHWs then gave pregnant women a specially designed information leaflet containing the same set of key messages as covered during the counselling, including pictures illustrating pregnancy-complication danger signs. We intended this leaflet to act as a reference guide for women to use during their antenatal period. CHWs also recorded information about the birth-preparedness and complication-readiness decisions made by pregnant woman and their families in a birth plan form for the trial. Then in the SBP and travel voucher arm, in addition to the structured birth planning, CHWs also gave pregnant women an EmONC travel voucher. This allowed them to get a cash reimbursement of eight US dollars (equivalent to 800 Pakistani Rupees) for transport costs if they accessed EmONC services from a MNCH Programme endorsed comprehensive EmONC facility (in practice a district hospital). We therefore hoped this would encourage women and family members to seek EmONC when needed without hesitation due to concerns over transportation costs.

In the control arm, CHWs continued to give health education to pregnant women following their existing routine practices. This may have included information related to birth-preparedness and complication-readiness, but this would not have been provided in a structured way as CHWs were trained to do in the intervention arm, and CHWs in the control arm had no access to any of the materials developed for the structure birth planning component of the intervention (i.e. the directory, flipchart and information leaflet). Therefore, any such information would have been provided ad-hoc at the CHW’s discretion and mixed with the other routine antenatal and postnatal care information provided. More generally, in the control arm the CHWs, through their routine practice, focused on telling women and their family members on what to do, but not the specifics of how to do it. There were no modifications to the intervention during the trial.

### Data collection

CHWs collected all trial data on participating women during their routine antenatal and postnatal care visits (conducted at least monthly) using two tools. First, the routine maternal and child health (MCH) record cards, which are used by CHWs to monitor women and children’s health when visiting women’s homes, and are also used by pregnant women when visiting a health facility. For the trial the MCH cards used by CHWs in all arms were modified slightly by the addition of a section to record pregnancy outcomes. The MCH record cards are known to be well maintained [4]. Second, CHWs in the intervention arms recorded the details of any key decisions, including plans for type of birth attendant and/or delivery location, made during the structured birth planning sessions on a separate card attached to the MCH card specifically for the trial. During CHWs’ routine monthly meetings at their RHC the MCH cards and additional forms were collected and checked for completeness and accuracy by their supervisors. Following routine MNCH Programme processes, lady health supervisors cross-checked and validated samples of MCH card records during random community visits. To further ensure high quality data researchers also conducted random telephone checks with registered women to validate collected data.

Researchers then took photographic copies of all records at regular intervals, which they then entered in batches into a Microsoft Access 2010 database with appropriate data validation checks. To further ensure data quality, they randomly checked 10% of the data in each batch against original records, re-entering that batch of data if the error rate exceeded 0.3%.

### Outcomes

We defined two co-primary outcomes, recorded at the level of the individual. We defined skilled birth attendance as a binary indicator, where births attended by either a doctor or a midwife (either a lady health visitor or a community midwife) were classed as skilled, and those attended by a traditional birth attendant were classed as not skilled. We defined neonatal mortality as a binary indicator, where a pregnancy resulting in either a still birth or a live birth where the child subsequently died from any cause within 28 days of delivery was classed as neonatal mortality. We chose these outcomes to evaluate the effectiveness of the interventions at 1) increasing the use of safe birthing (skilled attendance) services by pregnant women, and 2) reducing neonatal mortality.

To evaluate whether the intervention also influenced maternal health outcomes from pregnancy we also defined a secondary outcome of maternal mortality as a binary indicator of whether a pregnant woman died of any cause within 42 days of giving birth or survived past this point. To evaluate whether the intervention influenced birth attendance by specific types of attendant, subsequent to the protocol we also defined four secondary binary outcomes indicating whether or not a birth was attended by 1) a doctor, 2) a lady health visitor, 3) a community midwife or 4) a traditional birth attendant. An additional co-primary outcome of usage of EmONC facilities was dropped subsequent to the protocol because we were not able to collect the required data.

### Statistical analysis

We based the sample size on the neonatal mortality rate and assumed that any change in this rate would be much smaller than any change in the skilled birth attendance rate. We assumed a base rate of 58 neonatal deaths per 1,000 live births based on the 2007 Pakistan Demographic Health Survey [5]. We aimed to be able to detect a reduction in the neonatal mortality rate of at least 25% (to 43 or fewer deaths per 1,000 live births), in either of the intervention arms when compared to the control arm [6], with 80% power at the 5% significance level. Assuming an intra-cluster correlation coefficient of 0.0012, equal cluster sizes and a loss to follow-up rate of 20%, we required 21 clusters (7 per arm) with an average of at least 1,395 participants per cluster (i.e. 9,765 participants per arm and 29,295 participants overall).

As all our outcomes were binary we analysed them using generalised linear mixed-effects models with binomial errors and logit links, fitted using the LME4 package [7] in R version 3.3.1 [8], and accounted for the clustered design via random effects [9]. Using these models we made inferences at the individual level, comparing all outcomes between each intervention arm and the control arm, by estimating crude and adjusted odds ratios, their 95% confidence intervals and associated p-values based on two-sided hypothesis tests (using Wald tests) at the 5% level. For our covariate-adjusted analyses we adjusted for district (Chiniot, Jhang and Khanewal) and location (urban/rural) of birth due to the observed sizable imbalance in these variables between arms. For our co-primary outcomes we accounted for multiple hypothesis testing (two outcomes and two treatment comparisons per outcome) by separately adjusting the crude and covariate-adjusted p-values for the treatment effects using the Benjamini & Hochberg method [10]. We analysed all participants and clusters according to the treatment arm that they were originally allocated to, and as levels of missing data were so low we analysed completed cases only (i.e. for the crude analyses we excluded women who had missing outcome data, and for the covariate adjusted analyses we excluded women who had missing outcome or covariate data).

### Registration and ethics

The trial was registered with Current Controlled Trials (ISRCTN: 86264432. DOI 10.1186/ISRCTN86264432). Approvals were obtained from the National Bioethics Committee (NBC) Pakistan (Ref No.4-87/10NBC-39/RDC/487).

## Results

We randomised seven clusters to each arm, and lost none during the study. Between November 2011-July 2012, across all clusters, 2,152 CHWs identified and recruited a total of 59,505 pregnant women into the study, far exceeding the targeted sample size. Follow-up of pregnancy outcomes occurred between August-May 2012. Cluster sizes were highly variable within arms, and there was more cluster-size variation in the control arm compared to the intervention arms (SBP and travel voucher arm range = 1,871-6,818, SBP alone arm range = 1,035-4,953, control arm range = 150-8,009). However, the proportion of all participants enrolled into each arm was broadly comparable (Table 2). Loss to follow-up was minimal and substantially lower than the 20% expected, but was relatively greater in both intervention arms compared to the control arm (0.7% [157/21,371] in SBP and travel voucher arm, 0.7% [142/19,267] in SBP alone arm, and 0.3% [55/18,867] in the control arm; see Fig 1 for participant and cluster flow through the trial).

**Fig. 1.**
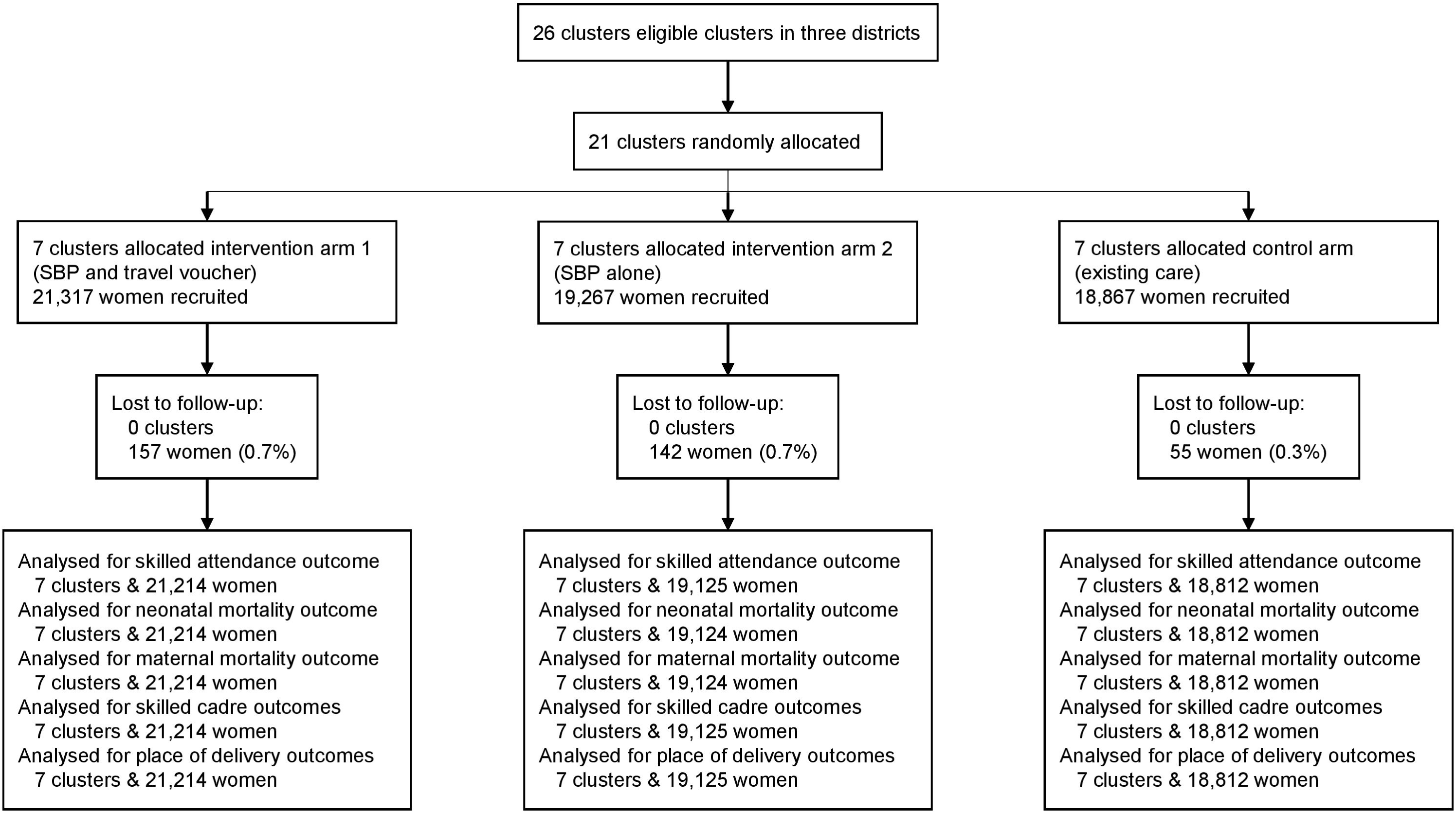
Trial flow diagram.

**Table 2.** Characteristics of pregnant women and clusters.

|  | Control<br>(N=18,867) | Arm 1 (SBP + travel voucher)<br>(N=21,371) | Arm 2 (SBP alone)<br>(N=19,267) |
| --- | --- | --- | --- |
| Age |  |  |  |
| <18 | 262 (1.4%) | 249 (1.2%) | 290 (1.6%) |
| 18-35 | 16,019 (87.7%) | 17,745 (87.9%) | 15,779 (84.4%) |
| >35 | 1,991 (10.9%) | 2,202 (10.9%) | 2,633 (14.1%) |
| Missing | 540 (2.9%) | 1,018 (4.8) | 423 (2.2) |
| Education (years) |  |  |  |
| Illiterate | 6,614 (60%) | 7,154 (52.1%) | 5,582 (51.7%) |
| 1-5 | 2,208 (20%) | 3,039 (22.1%) | 1,602 (14.8%) |
| 6-10 | 1,857 (9.9%) | 3,240 (15.3%) | 3,041 (15.9%) |
| 11+ | 339 (1.8%) | 309 (1.5%) | 569 (3%) |
| Missing | 7,794 (41.4%) | 7,472 (35.2%) | 8,331 (43.6%) |
| No. previous deliveries | 2.4 (1.8) | 2.4 (1.6) | 2.4 (1.5) |
| Missing | 1,593 (8.4%) | 854 (4%) | 2,560 (13.3%) |
| No. previous pregnancies | 2.9 (1.8) | 2.9 (1.6) | 2.93 (1.6) |
| Missing | 411 (2.2%) | 139 (0.7%) | 407 (2.1%) |
| No. previous live children | 2.3 (1.8) | 2.3 (1.6) | 2.3 (1.5) |
| Missing | 857 (4.5%) | 307 (1.4%) | 776 (4%) |
| Chronic disease |  |  |  |
| Yes | 287 (1.6%) | 458 (2.6%) | 721 (3.9%) |
| No | 17,420 (98.4%) | 17,085 (97.4%) | 17,896 (96.1%) |
| Missing | 1,105 (5.9%) | 3,671 (17.3%) | 508 (2.7%) |
| History of still birth |  |  |  |
| Yes | 572 (3.1%) | 638 (3.4%) | 543 (2.9%) |
| No | 17,835 (96.9%) | 17,960 (96.6%) | 18,340 (97.1%) |
| Missing | 405 (2.2%) | 2,616 (12.3%) | 242 (1.3%) |
| History of C-section |  |  |  |
| Yes | 1,204 (6.6%) | 590 (3.2%) | 1,340 (7.2%) |
| No | 17,171 (93.4%) | 17,926 (96.8%) | 17,404 (92.9%) |
| Missing | 437 (2.3%) | 2,698 (12.7%) | 381 (2%) |
| Birth district |  |  |  |
| Chiniot | 333 (0.6%) | 0 (0%) | 5,056 (8.5%) |
| Jhang | 3,573 (6%) | 11,645 (19.7%) | 3,089 (5.2%) |
| Khanewal | 14,906 (79.2%) | 9,569 (45.1%) | 10,980 (57.4%) |
| Missing | 55 (0.3%) | 157 (0.7%) | 142 (0.7%) |
| Birth location context |  |  |  |
| Urban | 2,313 (12.3%) | 2,015 (9.5%) | 1,031 (5.4%) |
| Rural | 16,499 (87.7%) | 19,199 (90.5%) | 18,094 (94.6%) |
| Participants lost to follow-up | 55 (0.3%) | 157 (0.7%) | 142 (0.7%) |
| Number of clusters | 7 | 7 | 7 |
| Clusters per district |  |  |  |
| Chiniot | 3 (37.5%) | 0 (0%) | 2 (28.6%) |
| Jhang | 3 (37.5%) | 5 (71.4%) | 2 (28.6%) |
| Khanewal | 2 (25%) | 2 (28.6%) | 3 (42.9%) |
| Births per cluster | 2,687 (150-8,009) | 3,030 (1,871-6,818) | 2,732 (1,035-4,953) |
| Number of facilities | 58 | 49 | 55 |
| Facilities per cluster | 8.3 (1-20) | 7 (4-12) | 7.9 (4-15) |

Abbreviations: SBP = structured birth planning. Data are n, n (%), mean (SD) or mean (lower range-upper range). All statistics are based on complete cases, with levels of missing data given.

Most women were aged between 18 and 35 and had previously had at least two children (Table 2). There were no substantial imbalances between arms in terms of having a history of still birth or having any chronic disease during the trial, but there was a moderately lower proportion of women who had a history of C-section in the SBP alone arm compared to either the SBP and travel voucher arm or the control arm (Table 2). There was also a moderately higher proportion of women who gave birth in urban areas in the SBP alone arm compared to either the SBP and travel voucher arm or the control arm, and there were also some substantial imbalances between arms in the distribution of participants between districts due to the randomisation (Table 2).

Both the crude and adjusted results showed that skilled birth attendance was not significantly different in the SBP and travel voucher arm compared to the control arm (Table 3). However, while the crude results indicated no significant difference in skilled birth attendance between the SBP alone arm and the control arm, once we adjusted for covariates there was a significantly higher level of skilled attendance in the SBP alone arm compared to the control arm, although the difference was modest (Table 3). Both the crude and adjusted results showed that neonatal mortality was significantly and substantially higher in the SBP and travel voucher arm (45 per 1,000 live births) compared to the control arm (13.4 per 1,000 live births), but was not significantly different in the SBP alone arm (16.2 1,000 live births) compared to the control arm (Table 3). Both crude and adjusted results also showed no significant difference in maternal mortality between either of the intervention arms compared to the control arm (Table 4).

**Table 3.** Crude and adjusted co-primary outcome results.

| Outcome | Control<br>(N=18,812) | SBP & travel voucher<br>(N=21,214) | SBP alone<br>(N=19,125) |
| --- | --- | --- | --- |
| Skilled birth attendance |  |  |  |
| n (%) | 12,045 (64%) | 15,335 (72.3%) | 14,944 (78.1%) |
| OR (95% CI); P-value | - | 1.15 (0.6, 2.17); 0.67 | 2.11 (1.03, 4.3); 0.08 |
| aOR (95% CI); P-value | - | 1.56 (0.93, 2.62); 0.21 | 2.16 (1.29, 3.62); 0.007 |
| Neonatal mortality (0-28 days) |  |  |  |
| Rate (/1,000 live births) | 13.4 | 45 | 16.2 |
| OR (95% CI); P-value | - | 2.95 (1.47, 5.93); 0.01 | 0.85 (0.41, 1.76); 0.68 |
| aOR (95% CI); P-value | - | 2.32 (1.48, 3.63); <0.001 | 1.36 (0.84, 2.21); 0.21 |

Abbreviations: SBP = structured birth planning. OR = unadjusted odds ratio. aOR = adjusted odds ratio.
Skilled birth attendance = births attended by either a doctor, lady health visitor or community midwife. The crude and adjusted sets of p-values are each separately adjusted for the multiple hypothesis testing resulting from having two co-primary outcomes. Treatment arm effects are obtained from multilevel logistic regression models with either treatment arm as the only independent variable (ORs), or with treatment arm, district and location of birth (urban/rural) as independent variables (aORs). Neonatal mortality crude/adjusted ICC = 0.12/0.04. Skilled birth attendance crude/adjusted ICC = 0.13/0.07.

**Table 4.** Crude and adjusted secondary outcome results.

| Outcome | Control<br>(N=18,812) | SBP & travel voucher<br>(N=21,214) | SBP alone<br>(N=19,125) |
| --- | --- | --- | --- |
| Maternal mortality (0-42 days) |  |  |  |
| n (%) | 24 (0.13%) | 46 (0.22%) | 35 (0.18%) |
| OR (95% CI); P-value | - | 1.61 (0.77, 3.39); 0.21 | 1.31 (0.61, 2.83); 0.49 |
| aOR (95% CI); P-value | - | 1.31 (0.72, 2.37); 0.38 | 1.58 (0.85, 2.95); 0.15 |
| LHV birth attendance |  |  |  |
| n (%) | 4,878 (25.9%) | 9,569 (45.1%) | 8,545 (44.7%) |
| OR (95% CI); P-value | - | 1.88 (1.36, 2.58); <0.001 | 1.79 (1.3, 2.45); <0.001 |
| aOR (95% CI); P-value | - | 1.99 (1.44, 2.74); <0.001 | 1.75 (1.29, 2.38); <0.001 |
| Doctor birth attendance |  |  |  |
| n (%) | 6,174 (32.8%) | 3,748 (17.7%) | 4,991 (26.1%) |
| OR (95% CI); P-value | - | 0.40 (0.27, 0.61); <0.001 | 0.76 (0.51, 1.13); 0.18 |
| aOR (95% CI); P-value | - | 0.47 (0.32, 0.69); <0.001 | 0.74 (0.51, 1.06); 0.1 |
| CMW birth attendance |  |  |  |
| n (%) | 993 (5.3%) | 2,018 (9.5%) | 1,408 (7.4%) |
| OR (95% CI); P-value | - | 1.31 (0.68, 2.53); 0.42 | 0.97 (0.5, 1.88); 0.93 |
| aOR (95% CI); P-value | - | 1.13 (0.76, 1.69); 0.54 | 1.13 (0.77, 1.67); 0.52 |
| Traditional birth attendant<br>(unskilled) birth attendance |  |  |  |
| n (%) | 6,767 (36%) | 5,879 (27.7%) | 4,181 (21.9%) |
| OR (95% CI); P-value | - | 0.87 (0.42, 1.8); 0.71 | 0.47 (0.23, 0.97); 0.04 |
| aOR (95% CI); P-value | - | 0.64 (0.37, 1.12); 0.12 | 0.46 (0.27, 0.78); 0.004 |
| Home delivery |  |  |  |
| n (%) | 7,685 (40.9%) | 7,078 (33.4%) | 4,948 (25.9%) |
| OR (95% CI); P-value | - | 0.99 (0.51, 1.94); 0.99 | 0.5 (0.24, 1.05); 0.07 |
| aOR (95% CI); P-value | - | 0.67 (0.41, 1.1); 0.11 | 0.49 (0.31, 0.8); 0.003 |
| CMW's home delivery |  |  |  |
| n (%) | 646 (3.4%) | 1,277 (6%) | 1,091 (5.7%) |
| OR (95% CI); P-value | - | 1.34 (0.55, 3.31); 0.52 | 1 (0.41, 2.47); 0.99 |
| aOR (95% CI); P-value | - | 1.23 (0.64, 2.34); 0.54 | 1.17 (0.63, 2.19); 0.62 |
| Health facility delivery |  |  |  |
| n (%) | 2,633 (14%) | 5,894 (27.8%) | 5,842 (30.5%) |
| OR (95% CI); P-value | - | 1.4 (0.72, 2.73); 0.32 | 1.6 (0.86, 3); 0.14 |
| aOR (95% CI); P-value | - | 1.64 (1.07, 2.51); 0.02 | 1.81 (1.21, 2.7); 0.004 |
| Hospital delivery |  |  |  |
| n (%) | 7,848 (41.7%) | 6,965 (32.8%) | 7,244 (37.9%) |
| OR (95% CI); P-value | - | 0.69 (0.43, 1.1); 0.12 | 0.84 (0.5, 1.42); 0.52 |
| aOR (95% CI); P-value | - | 0.8 (0.54, 1.18); 0.26 | 0.75 (0.52, 1.09); 0.13 |

Abbreviations: SBP = structured birth planning, LHV = lady health visitor, CMW = community midwife. OR = unadjusted odds ratio. aOR = adjusted odds ratio.
Treatment arm effects are obtained from multilevel logistic regression models with either treatment arm as the only independent variable (ORs), or with treatment arm, district and location of birth (urban/rural) as independent variables (aORs).

In our exploratory analyses looking at changes in birth attendance and location we found that attendance by lady health visitors was significantly and substantially higher in both intervention arms compared to the control arm (Table 4). The majority of lady health visitor-led births occurred in health facilities, which include RHCs, BHUs and private clinics, as well as hospitals, which include both public (district and sub-district hospitals) and private hospitals (Table 5). We also found that birth attendance by doctors was significantly and substantially lower in the SBP and travel voucher arm compared to the control arm, but was not significantly different between the SBP alone arm and the control arm (Table 4). The majority of doctor-led births occurred in hospitals (Table 5). However, there was no significant difference in birth attendance by community midwives between either of the intervention arms and the control arm (Table 4), and most community midwife-led births occurred in community midwives’ homes or women’s homes (Table 5). We also found that traditional birth attendance was significantly, albeit only moderately, lower in the SBP alone arm compared to the control arm, but was not significantly different between the SBP and travel voucher arm and the control arm (Table 4). The vast majority of traditional birth attendant-led births occurred in women’s homes (Table 5). For all these outcomes there were no substantive differences between the crude and adjusted results.

**Table 5.** Neonatal (age 0-28 days) mortality by treatment arm, birth attendant and place of delivery.

| Birth location & attendant | Control |  | SBP & travel voucher |  | SBP alone |  |
| --- | --- | --- | --- | --- | --- | --- |
|  | Births<sup>a</sup> | NNM<sup>b</sup> | Births<sup>a</sup> | NNM<sup>b</sup> | Births<sup>a</sup> | NNM<sup>b</sup> |
| Women's home –LHV | 874 (4.6%) | 10 (1.1%) | 790 (3.7%) | 29 (3.7%) | 609 (3.2%) | 7 (1.1%) |
| Women's home – CMW | 315 (1.7%) | 2 (0.6%) | 649 (3.1%) | 21 (3.2%) | 251 (1.3%) | 2 (0.8%) |
| Women's home –TBA | 6,496 (34.5%) | 59 (0.9%) | 5,639 (26.6%) | 180 (3.2%) | 4,088 (21.4%) | 61 (1.5%) |
| CMW's home – LHV | 56 (0.3%) | 0 (0%) | 149 (0.7%) | 7 (4.7%) | 145 (0.8%) | 11 (7.6%) |
| CMW's home – CMW | 478 (2.5%) | 9 (1.9%) | 1,032 (4.9%) | 51 (4.9%) | 916 (4.8%) | 22 (2.4%) |
| CMW's home –TBA | 112 (0.6%) | 0 (0%) | 96 (0.5%) | 4 (4.2%) | 30 (0.2%) | 1 (3.3%) |
| Health facility - Doctor | 377 (2%) | 18 (4.8%) | 497 (2.3%) | 89 (17.9%) | 357 (1.9%) | 24 (6.7%) |
| Health facility – LHV | 2,125 (11.3%) | 19 (0.9%) | 5,103 (24.1%) | 249 (4.9%) | 5,245 (27.4%) | 43 (0.8%) |
| Health facility – CMW | 74 (0.4%) | 1 (1.4%) | 210 (1%) | 33 (15.7%) | 196 (1%) | 28 (14.3%) |
| Health facility –TBA | 57 (0.3%) | 1 (1.8%) | 84 (0.4%) | 10 (11.9%) | 44 (0.2%) | 2 (4.5%) |
| Hospital – Doctor | 5,797 (30.8%) | 100 (1.7%) | 3,251 (15.3%) | 160 (4.9%) | 4,634 (24.2%) | 79 (1.7%) |
| Hospital – LHV | 1,823 (9.7%) | 31 (1.7%) | 3,527 (16.6%) | 115 (3.3%) | 2,546 (13.3%) | 29 (1.1%) |
| Hospital – CMW | 126 (0.7%) | 1 (0.8%) | 127 (0.6%) | 2 (1.6%) | 45 (0.2%) | 1 (2.2%) |
| Hospital –TBA | 102 (0.5%) | 2 (2%) | 60 (0.3%) | 4 (6.7%) | 19 (0.1%) | 0 (0%) |
| All | 18,812 | 253 | 21,214 | 954 | 19,125 | 310 |

Abbreviations: SBP = structured birth planning, NNM rate = neonatal (0-28 days) mortality percentage, LHV = lady health visitor, CMW = community midwife, TBA = traditional birth attendant (Dai).
Data are n (%).
<sup>a</sup>Birth percentages are calculated based on the number of births for each category (e.g. "Women's home – LHV") divided by the total number of births for that arm. <sup>b</sup>Neonatal mortality percentages are specific to each category, i.e. they are calculated based on the number of neonatal deaths for each category within each arm (e.g. neonatal deaths for control arm category "Women's home – LHV") divided by the total number of births for that category within that arm.
Health facility = RHC or private clinic, including those run by unqualified/semi-qualified staff. Hospital = public sub-district or district hospitals or private hospitals.
Note: there were no deliveries by doctors at women's homes or at CMW's homes.

In terms of birth location we found that levels of home deliveries were not significantly different between the SBP and travel voucher arm compared to the control arm (both crude and adjusted results), but that there was a significant (although only after covariate adjustment) and sizably lower level in the SBP alone arm compared to the control (Table 4). There were no significant differences in the level of births at community midwives’ homes between either of the intervention arms and the control arm (Table 4). There was a much larger proportion of births in health facilities in both intervention arms compared to the control arm, although the difference was only statistically significant after adjustment for covariates (Table 4). Finally, there were no significant differences in the level of births at hospitals between either of the intervention arms and the control arm, even after covariate adjustment (Table 4).

There were some clear differences between treatment arms in the distribution of births across the possible birth location and birth attendant type combinations (Table 5). The biggest differences in absolute terms were the much higher numbers of births at health facilities attended by lady health visitors in both intervention arms compared to the control arm, the much lower number of births at hospitals attended by doctors in the SBP and travel voucher arm, and to a lesser extent the SBP alone arm, compared to the control arm, and the much lower number of births at women’s homes attended by traditional birth attendants in the SBP alone arm, and to a lesser extent in the SBP and travel voucher arm, compared to the control arm. However, these differences did not clearly relate to the observed differences in neonatal mortality between the SBP and travel voucher arm and the control arm. In terms of absolute numbers, most of the excess neonatal mortality in the SBP and travel voucher arm compared to the control arm occurred in those location-attendant combinations where most births occurred, particularly (in descending order) births at health facilities attended by lady health visitors, births at women’s homes attended by traditional birth attendants, births at hospitals attended by doctors and lady health visitors, and births at health facilities attended by doctors (Table 5). However, in both relative and absolute terms neonatal mortality rates were higher, often substantially so, across all location-attendant combinations in the SBP and travel voucher arm compared to the same location-attendant combinations in the control arm (and often the SBP alone arm), indicating likely universally poorer neonatal outcomes in this arm compared to the control arm and SBP alone arm irrespective of birth location and birth attendant type.

## Discussion

We assumed that the birth-preparedness planning component of our intervention would increase the proportion of women giving birth with a skilled attendant, and that the complication-readiness planning and information leaflet component would increase the frequency and speed with which women (potentially aided by family members) seek EmONC when needed. We also expected that the travel voucher would further support more frequent and rapid EmONC seeking behaviour by removing/reducing financial barriers to accessing EmONC. We also predicted that together these interventions would lead to safer birthing practices, ultimately reducing the neonatal mortality rate and the maternal mortality rate. Interestingly though, our data showed that only 1.3% of women in the SBP and travel voucher arm who had caesarean sections (i.e. accessed EmONC) used a travel voucher, showing that the travel vouchers were actually very rarely used by women when they could have been. Therefore, in practice both intervention arms appear to have operated in a very similar way, given that the only difference between them was whether the travel voucher was provided by CHWs to women or not. Overall the trial successfully recruited a far larger number of women than expected, and successfully implemented the interventions in each arm.

Contrary to our expectations we only found evidence for a modestly higher proportion of skilled birth attendance in the SBP alone arm compared to the control arm, and no difference between the SBP and travel voucher arm compared to the control arm. Looking at the intervention’s effects on attendance by specific types of skilled attendant may help explain these results. While we found that the proportion of births attended by lady health visitors were much higher in both intervention arms compared to the control arm (as expected due to the birth-preparedness planning components of the intervention), we also found that the proportion of births attended by doctors was much lower in the SBP and travel voucher arm, but not the SBP alone arm, compared to the control arm. It is not clear why this was the case, but it is unlikely to be due to the SBP intervention or we would expect the same changes in both intervention arms. We also found no differences in the proportion of births attended by community midwives between either of the intervention arms and the control arm. Therefore, compared to the control arm it appears that the higher proportion of overall skilled birth attendance in the SBP alone arm was primarily driven by more births being attended by lady health visitors. However, while the SBP and travel voucher arm also displayed a higher proportion of births attended by lady health visitors compared to the control arm, any difference in the overall proportion of skilled birth attendance compared to the control arm was cancelled out by the much lower proportion of births attended by doctors (the other skilled cadre of community midwives had little effect either way given the relatively low proportion of births attended by them).

Given the similarity in practice of the two intervention arms though it is not clear why births attended by doctors were so much lower in the SBP and travel voucher arm compared to the control arm, but not in the SBP alone arm compared to the control arm. As expected due to the effects of the birth-preparedness intervention encouraging women to use skilled attendants, births attended by traditional (unskilled) attendants were lower in the SBP alone arm compared to the control arm, but it is again unclear why they were not also lower in the SBP and travel voucher arm compared to the control arm. Therefore, with hindsight the overall skilled attendance outcome appears to have been a crude measure of the effects of the interventions on skilled attendance, which failed to capture the likely shift in birth attendance away from traditional birth attendants and, to a lesser and more varied extent, doctors, in favour of lady health visitors.

Unexpectedly we found a much higher rate of neonatal mortality in the SBP and travel voucher arm compared to the control arm, which was very concerning, and no difference between the SBP alone arm and the control arm. The cause of these results is not clear, but a number of factors may be involved. First, when compared to the neonatal mortality rates estimated by the most recent Pakistan Demographic and Health [5] for Pakistan as a whole between 2008-2012 (55 per 1,000 live births), or for the Punjab province alone between 2003-2012 (63 per 1,000 live births), or even for the highest PDHS wealth quintile (34 per 1,000 live births), the estimated neonatal mortality rates for women in rural communities in the control (13.4 per 1,000 live births) and SBP alone arms (16.2 per 1,000 live births) do not appear to be realistic. It was not feasible for researchers to verify neonatal mortality or other outcomes with women. However, we used routine CHW MNCH card data collection processes that have previously been evaluated as robust [4], along with robust data quality assurance processes during data collection, collation and data entry. Despite this it appears likely that there was systematic underreporting of neonatal mortality in the control arm and the SBP alone arm. Unfortunately at present it is not clear why this apparent underreporting occurred, but investigating this issue and the quality of the MNCH Programme’s routine data (on birth outcomes in particular) is therefore a top priority for the next programme evaluation that is due to begin shortly and will involve members of this study.

Given that the data collection and verification processes were identical in each arm, and as far as we could tell operated to the same degree of quality in all arms, and given that in practice both intervention arms operated in essentially the same way it is unclear why this apparent underreporting only affected the control and SBP alone arms though. One possible explanation is that as a cluster randomised controlled trial with relatively few clusters (just 7 per arm), the simple randomisation process used may have failed to evenly distribute clusters with underreporting issues between the treatment and control arms, with more ending up in the SBP and travel voucher arm. It is also possible that more clusters with poorer underlying neonatal outcomes were allocated to the SBP and travel voucher arm via the simple randomisation process, which may also have contributed to the observed neonatal mortality results in this arm. This is an important trial limitation, and using a more sophisticated form of restricted/stratified randomisation informed by relevant data could have reduced any such bias. In addition, although it was not possible due to operational and cost limitations, having baseline data on neonatal mortality would have also allowed us to control for any imbalances in neonatal mortality rate reporting and/or underlying levels between arms.

In addition, the observed intra-cluster correlation coefficient for neonatal mortality was far higher than expected in our sample size calculation. This indicates that there was an unexpectedly high amount of variation in neonatal mortality outcomes at the cluster level, which suggests some cluster-level factors affected the underreporting of neonatal mortality. It also indicates that the cluster randomisation would have had more ability to influence the results by generating imbalances in levels of neonatal mortality reporting and/or underlying neonatal mortality rates between arms.

However, despite the likely issues with the neonatal mortality data it cannot be assumed that the higher rate of neonatal mortality in the SBP and travel voucher arm is completely spurious. This is because poor quality midwifery care has been identified as an issue by Pakistani Punjab provincial health experts (including from the MNCH and LHW Programmes, the district offer, district hospital and midwifery school) who have reviewed the results of the study. Specifically they highlighted concerns about poor existing practices by some midwifes, particularly by lady health visitors, who were responsible for the delivery of a far higher proportion of births in both intervention arms compared to the controls arms. For example, in Pakistan midwives (lady health visitors and community midwives) do not generally use partographs to track the progress of labour, which is a potential cause of delayed recognition of the need for EmONC referral. Lady health visitors may also use inappropriate emergency delivery procedures such as forceps, rather than the safer alternative ventouse vacuum-assisted procedure. It is therefore possible that increasing birth attendance by lady health visitors may have exposed more women to out of date practices and introduced delays in accessing needed EmONC, potentially more so than compared to home deliveries by traditional (unskilled) birth attendants who may be more likely to recognise that they do not have the skills to cope when there are complications and so refer women more rapidly to hospital/doctors. Furthermore, current postnatal care practices also seem inadequate, and may contribute to the failure to reduce mortality. In addition, the current management support arrangements lack regular in-service training and quality control procedures of midwives or other skilled providers in public or private clinics. The MNCH Programme has been focussed on expanding training of CMWs, and there is a lack of in-service training and quality control procedures for existing lady health visitors the existing midwives. All these issues will be further investigated in the next programme evaluation.

It is difficult to compare our data with other skilled birth attendance studies. This is due to differences in reporting accuracy, and because of differences in components of intervention and varied settings. In our study, we did not train traditional birth attendants, nor train or monitor the quality of care provided by midwives.

An earlier trial (2003-2005) in Sindh province in Pakistan involving training of CHWs and traditional birth attendants in delivering antenatal and postnatal care did increase the use of skilled birth attendants and lower the neonatal mortality rate [11]. An intervention study in Balochistan also increased the proportion of mothers with complications getting timely and appropriate help, improved neonatal care, and reduced maternal and neonatal mortality [12]. This intervention involved training traditional birth attendants, educating pregnant women and their husbands, improving the transport and communication system for accessing EmONC, and strengthening hospital EmONC services. These interventions in Pakistan had some common elements of improving care practices, enhancing service uptake through education, and providing transport for EmONC. Though they also trained traditional birth attendants, and in two of them also strengthened hospital EmONC services. Overall the findings from these studies support the need for strengthening the quality of care provided by all birth attendants.

Finally, we did not find any effect of the interventions on maternal mortality rates, but this was not unexpected given that the trial was not powered to detect plausible changes in these rates, which are much lower than the plausible changes in neonatal mortality that the trial was powered for.

Therefore, our study provided evidence that a CHW intervention involving structured birth planning (more specifically counselling of women on birth-preparedness and complication-readiness) has the potential to improve women’s usage of skilled birth attendants in Pakistan, in particular lady health visitors. However, we were not able to clearly understand the effects of these changes on neonatal mortality, given the questions over the completeness of the recording by CHWs on the MNCH record card, including crucially the recording of birth outcomes. This issue will be addressed by the forthcoming Lady Health Worker (CHW) Programme evaluation. The MNCH Programme has also reviewed these results, appraised options and agreed on a plan to develop and implement quality procedures, performance management, and public-private partnership models for improved quality of midwifery care.

## Data Availability

All data produced in the present study are available upon reasonable request to the authors

## Acknowledgments

We acknowledge the National and Provincial MNCH programme and Directorate health services Punjab for their valuable inputs in developing a contextualized intervention and steering the design and conduct of intervention evaluation. We would like to acknowledge valuable inputs of three respective district health offices; and facility and community based healthcare staff in developing the contextualized intervention and implementing the trial. Lastly we would also like to acknowledge Save the Children for their valuable contribution in the overall trial design and implementation.

## References

1. Campbell OMR, Graham WJ, Lancet Maternal Survival S. Maternal survival 2 - Strategies for reducing maternal mortality: getting on with what works. Lancet. 2006;368(9543):1284–99. doi: 10.1016/s0140-6736(06)69381-1. PubMed PMID: WOS:000241058300036.

2. McPherson RA, Khadka N, Moore JM, Sharma M. Are birth-preparedness programmes effective? Results from a field trial in Siraha district, Nepal. Journal of Health Population and Nutrition. 2006;24(4):479–88. PubMed PMID: WOS:000246598400012.

3. Khan MA, Mirza S, Ahmed M, Rasheed A, Khan A, Walley J, et al. Making birthing safe for Pakistan women: a cluster randomized trial. Bmc Pregnancy and Childbirth. 2012;12. doi: 10.1186/1471-2393-12-67. PubMed PMID: WOS:000309116000001.

4. Mid-term Evaluation of the National Maternal and Child Health Programme in Pakistan: Findings and Recommendations. Islamabad, Pakistan: 2012. Accessed on 1.08.2017 from: http://www.trfpakistan.org/LinkClick.aspx?fileticket=0WOWUr-6vQI%3D&tabid=2618

5. National Institute of Population Studies (NIPS) [Pakistan] and ICF International. 2013. Pakistan Demographic and Health Survey 2012-13. Islamabad, Pakistan, and Calverton, Maryland, USA: NIPS and ICF International.

6. Lassi ZS, Bhutta ZA. Community-based intervention packages for reducing maternal and neonatal morbidity and mortality and improving neonatal outcomes. Cochrane Database of Systematic Reviews. 2015;(3). doi: 10.1002/14651858.CD007754.pub3. PubMed PMID: WOS:000352608900019.

7. Bates D, Machler M, Bolker BM, Walker SC. Fitting Linear Mixed-Effects Models Using lme4. Journal of Statistical Software. 2015;67(1):1–48. PubMed PMID: WOS:000365981400001.

8. R Development Core Team. R: A language and environment for statistical computing. Vienna, Austria: R Foundation for Statistical Computing; 2016.

9. Hayes R, Moulton L. Cluster randomised trials. London: Chapman & Hall/CRC; 2009.

10. Benjamini Y, Hochberg Y. Controlling the false discovery rate: a practical and powerful approach to multiple testing. Journal of the Royal Statistical Society Series B-Methodological. 1995;57(1):289–300. PubMed PMID: WOS:A1995QE45300017.

11. Bhutta ZA, Memon ZA, Soofi S, Salat MS, Cousens S, Martines J. Implementing community-based perinatal care: results from a pilot study in rural Pakistan. Bulletin of the World Health Organization. 2008;86(6):452–9. doi: 10.2471/blt.07.045849. PubMed PMID: WOS:000257100300012.

12. Midhet F, Becker S. Impact of community-based interventions on maternal and neonatal health indicators: Results from a community randomized trial in rural Balochistan, Pakistan. Reproductive Health. 2010;7. doi: 10.1186/1742-4755-7-30. PubMed PMID: WOS:000208607600012.

